# Challenges for non-technical implementation of digital proximity tracing: early experiences from Switzerland

**DOI:** 10.1101/2020.10.22.20218057

**Authors:** Viktor von Wyl

## Abstract

**Background:** Several countries have released digital proximity tracing (DPT) apps in addition to manual contact tracing (MCT) to combat the Sars-CoV-2 pandemic. The goal of DPT is to notify app users about proximity exposures to persons infected with Sars-CoV-2 so that they can self-quarantine. However, early press reports from Switzerland suggest multiple challenges for non-technical DPT implementation.

**Objective:** Using media articles published during the first three months after the DPT app launch to describe non-technical implementation challenges reported by different stakeholders and to map these reports to the four constructs of normalization process theory (NPT), a framework to develop and evaluate complex digital health interventions.

**Methods:** A Swiss media database was searched for articles on the Swiss DPT app (SwissCovid) published in German or French between 04.07.2020 and 03.10.2020. Topics were extracted manually from articles that were deemed pertinent in a structured process. Extracted topics were mapped to NPT constructs.

**Results:** Out of 94 articles deemed pertinent and selected for closer inspection, 38 provided unique information on implementation challenges. These challenges included unclear DPT benefits, which affected commitment and raised fears among different health system actors regarding resource competition with established pandemic mitigation measures. Moreover, media reports indicated process interface challenges such as delays or unclear responsibilities in the notification cascade, as well as misunderstandings and unmet communication needs from certain health system actors. Finally, some reports suggested misaligned incentives, not only for app usage by the public but also for process engagement by other actors in the app notification cascade. These challenges mapped well to the four constructs of NPT.

**Conclusions:** Early experiences from one of the earliest adopters of DPT indicate that non-technical implementation challenges warrant attention. The detected implementation challenges fit well into the framework of NPT, which seems well suited to guide the development and evaluation of complex DPT interventions.

## Background

To combat the Sars-CoV-2 pandemic, several countries employ digital proximity tracing (DPT) smartphone apps to complement manual contact tracing (MCT^).1,2^ DPT enables tracing of proximity contacts who could pose relevant infection risks if, for example, two persons were in close proximity of 1.5m or less for 15 minutes or more. If one of the proximity contacts tests positive for Sars-CoV-2, all other app users with relevant proximity within the window of infectivity are warned by the app.

Most countries that have released DPT apps have opted for a privacy-preserving, decentralized architecture.^1,2^ That is, proximity contacts are not sent to a central server but are stored and evaluated locally on smartphones. The only data sent to a central server are anonymous random identifiers of persons with a confirmed SARS-CoV-2 infection. These “infectious” identifiers are downloaded by all other users and compared against the locally stored identifiers of proximity encounters. If a match is found, users are notified and information on further steps is provided.

DPT has some distinct advantages over manual contact tracing (MCT) because it can warn exposed contacts much faster, simultaneously, and can reach contacts not personally known to the index case.^3^ By contrast, MCT identifies index case contacts through interviews, which is a labor-intensive process.^4^ Given that exposed contacts enter quarantine upon app notification, the speed advantage of DPT should also lead to a faster interruption of transmission chains. But to achieve the desired goals, DPT needs adequate embedding in a country’s overall test-trace-isolate-quarantine (TTIQ) response against the epidemic.^5,6^

Switzerland was one of the first countries to release its own DPT app (“SwissCovid” app) on June 25 based on decentralized, privacy-preserving proximity tracing architecture (DP-3T).^2^ The overall principle of the SwissCovid notification cascade is illustrated in Figure 1. Of note, the app notification sequence involves multiple actors, including the testing laboratory, the physician ordering the test, MCT (which also generates the “CovidCodes”; that is, the notification codes to be uploaded by the positive tested user), and an infoline that notified users are recommended to call. While the SwissCovid infoline is centralized, MCT is individually organized at the level of the 26 cantons. The operational lead of MCT resides with the cantonal physician. Testing laboratories are decentrally organized private or public institutions. Guidelines and reporting forms are in place to inform the Federal Office of Public Health and the responsible cantonal health authorities about each newly detected Sars-CoV-2 case.

**Figure 1:**
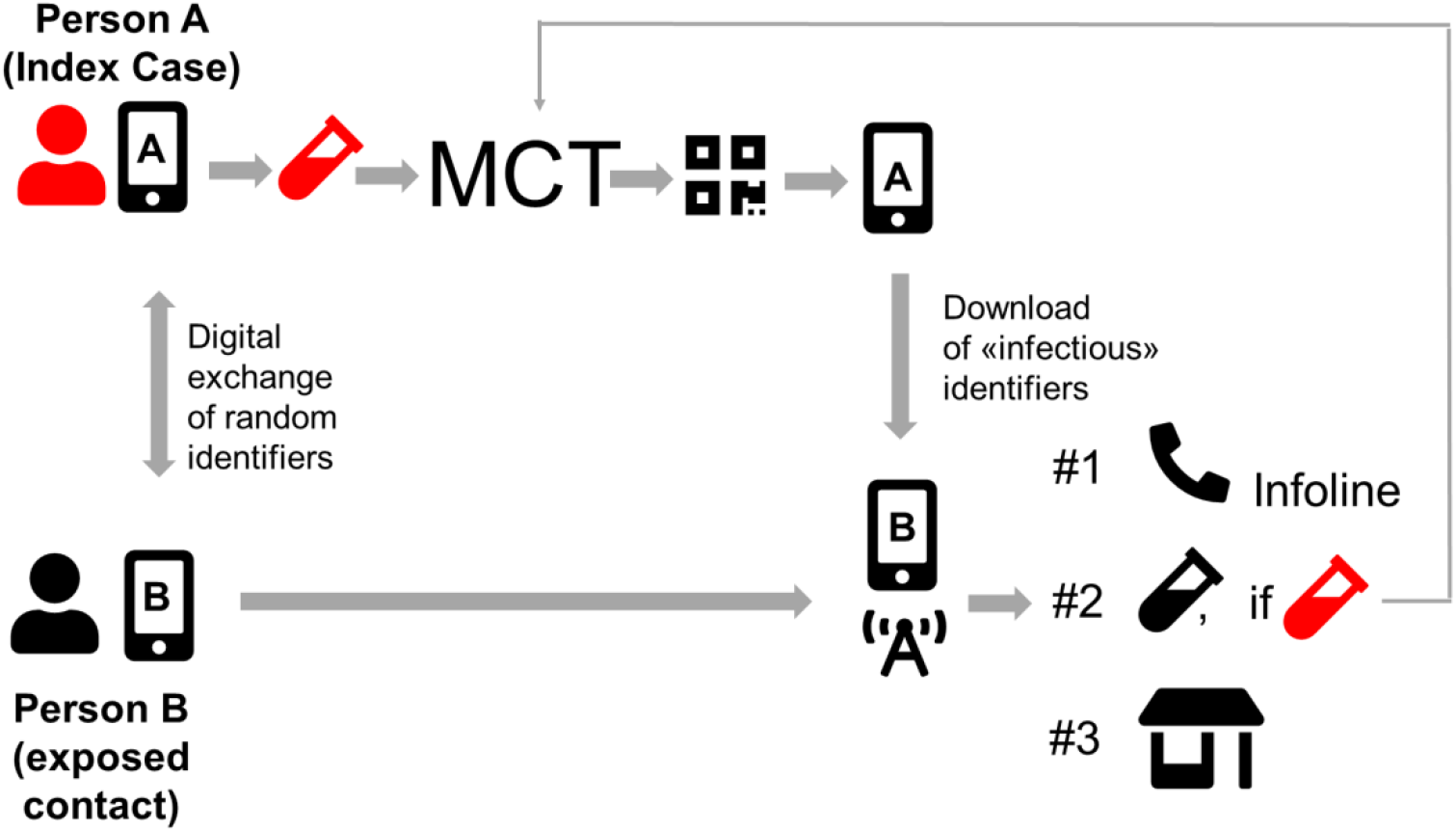
Steps in the notification cascade of digital proximity tracing. An infected person A gets tested positive for Sars-CoV-2, is referred to manual contact tracing (MCT) and receives and uploads a CovidCode to warn other app users. Person B was in close proximity and got infected. This person receives the app notification, upon which she has several options: calling an infoline (#1, recommended option), receiving a free test (#2), and/or staying home voluntarily (#3). Abbreviations: MCT, manual contact tracing.

App-notified users have several options on how to proceed. The app notification suggests calling an official infoline. The infoline informs users that they are eligible for a free test five days after exposure notification and inquires more specifically about the potential risk exposure. If indicated, the infoline recommends a voluntary quarantine (Figure 1, #1). By comparison, MCT can order and enforce mandatory quarantine, but with salary compensation for working persons. Furthermore, notified app users can also directly request a free PCR-test (Figure 1, #2) or voluntarily stay at home without calling the infoline (Figure 1, #3), in which case they would not appear in any statistics. In the first phase after the app launch, the infoline could not directly refer callers to the responsible cantonal authorities and manual contact tracing, but since September 2020 a respective agreement is in place.

Some of the notification cascade steps have relatively strict timelines. Laboratories and/or attending physicians must report positive PCR-test results to the responsible cantonal physician within two hours. The cantonal physician should generate the CovidCode and hand it over to the infected app user, who has 24 hours to enter the code (after which the code expires).

In implementation science, complex interventions are defined as “consisting of multiple behavioral, technological, and organizational components”.^7^ As illustrated by Figure 1, DPT fulfills this definition of complex public health interventions because it involves several steps from laboratory testing, communication of results, notification triggering, notification receipt, and quarantine. Consequently, the success of DPT hinges on an efficient cascade of notifications from PCR-positive test results to proximity contacts and involves app users, Sars-CoV-2 testing laboratories, health authorities, and possibly other actors. Therefore, seamless integration of DPT into broader pandemic mitigation measures (e.g. testing facilities) and high app user compliance with recommended measures (e.g. trigger notifications, entering quarantine) is crucial, especially in settings with voluntary DPT usage.

Early data from September 2020 about SwissCovid performance, both in terms of individual notification cascade steps and overall, paint a mixed picture. A recent report suggests proof-of-principle for the technical infrastructure of DPT.^8^ Specifically, the study reports on at least 60 persons who were tested after an app notification and who turned out to be PCR-positive for Sars-CoV-2. However, the same report also highlights some inefficiencies in the notification chain as described above. For example, the number of CovidCodes exceeds the number of entered codes by around 50%, which – in part – is the result of the voluntary nature of DPT: at each step, users can select whether or not to use the app and undertake the recommended steps, without fear of retributions. Along similar lines, a separate report examined uptake and reasons for non-use of the SwissCovid app.^9^ For example, higher monthly household income or being a non-smoker were associated with higher SwissCovid app uptake; whereas older age, lack of trust in health authorities, or having a non-Swiss nationality correlated with a lower uptake. Combined, both reports underscore the relevance of non-technical implementation aspects for optimal DPT functioning.^9^

Furthermore, the media coverage of SwissCovid during the first three months unearthed some organizational challenges and examples of cases where steps in the notification cascade were not handled optimally. This is not completely surprising given the novelty and complexity of the intervention. In other instances of complex intervention assessments, normalization process theory (NPT) has proven useful to systematically investigate the embedding of complex (digital) health interventions.^7,10^ NPT aims to explain and promote factors that normalize an intervention, that is, to make it part of routine practice. NPT is centered around four core constructs, which also reflect the flow of an intervention development from planning, stakeholder onboarding, and intervention execution to critical appraisal.^11^ Along this “life course” of an intervention, the *coherence* constructs describe how individual participants make sense of implementation, *cognitive participation* reflects the participants’ collective efforts to create commitment and engagement with the intervention, *collective action* refers to the execution of the intervention and describes the joint efforts of all actors to make it work, and *reflexive monitoring* refers to collective appraisals of risks and benefits, and developing improvements by all actors.^11^

This analysis aimed to systematically scrutinize media reporting on SwissCovid for statements and examples reflecting challenges to optimal intervention functioning. Furthermore, by mapping challenges to the NPT constructs, it was systematically assessed whether NPT is a useful approach to group early experiences of non-technical implementation of the SwissCovid app. The results contribute to the literature by exploring the utility of NPT as a reporting framework for DPT implementation and by summarizing important lessons from one of the first countries to nationally adopt DPT.

## Methods

The present analysis was informed by Swiss media reports. The Swissdox Essentials media database (www.swissdox.ch) was searched from July 4, 2020, to October 3, 2020. This database covers all Swiss print media and the most important online portals. Only entries in German or French were considered, which are spoken by 85% of Swiss inhabitants (https://www.bfs.admin.ch/bfs/en/home/statistics/population/languages-religions/languages.assetdetail.11887091.html). Using the search phrase “SwissCovid or (“Swiss Covid” AND app)", the Swissdox database was searched for unique print articles and online articles reflecting independent journalistic investigations. That is, reprinted articles or articles referring to other articles without adding information were excluded. Live ticker transcripts were also excluded from our search.

Eligible articles were pre-screened for whether they report on problems or inefficiencies of the SwissCovid app. In a second, more detailed search, duplicate articles were removed (also those just paraphrasing an earlier article), and topics were extracted based on the following, pre-defined list (informed by subject knowledge of the author): problems referring to (a lack of) communication, technical problems or confusion regarding the app, issues related to the effectiveness of the app, delays in receiving test results, delays in receiving CovidCodes, issues related to the infoline, lack of support from participants, or competition for strained resources by SwissCovid. The key issues regarding the SwissCovid app were summarized and grouped by the main author.

Finally, the key challenges reported in the press were contextualized and interpreted using the NPT questions developed by Murray et al.^10^ NPT was selected a priori as an assessment framework based on findings in the literature. The present study followed the guiding questions outlined by Murray et al.^10^ and considered the following stakeholders involved in the DPT notification cascade in Switzerland (henceforth also called “participants” to remain compatible with NPT terminology): app users; Sars-CoV-2 testing laboratories; cantonal health authorities and cantonal physicians who operate MCT; the FOPH, which is the product owner of the SwissCovid app; and the infoline, operated on behalf of the FOPH by a commercial telehealth company.

## Results

### Findings from the literature search

Figure 2 outlines the search process of the media database, which resulted in a total of 39 articles deemed relevant for the analysis. The key topics extracted from the selected articles are outlined in Table 1 (with raw data presented in supplementary table 1).

**Table 1:**
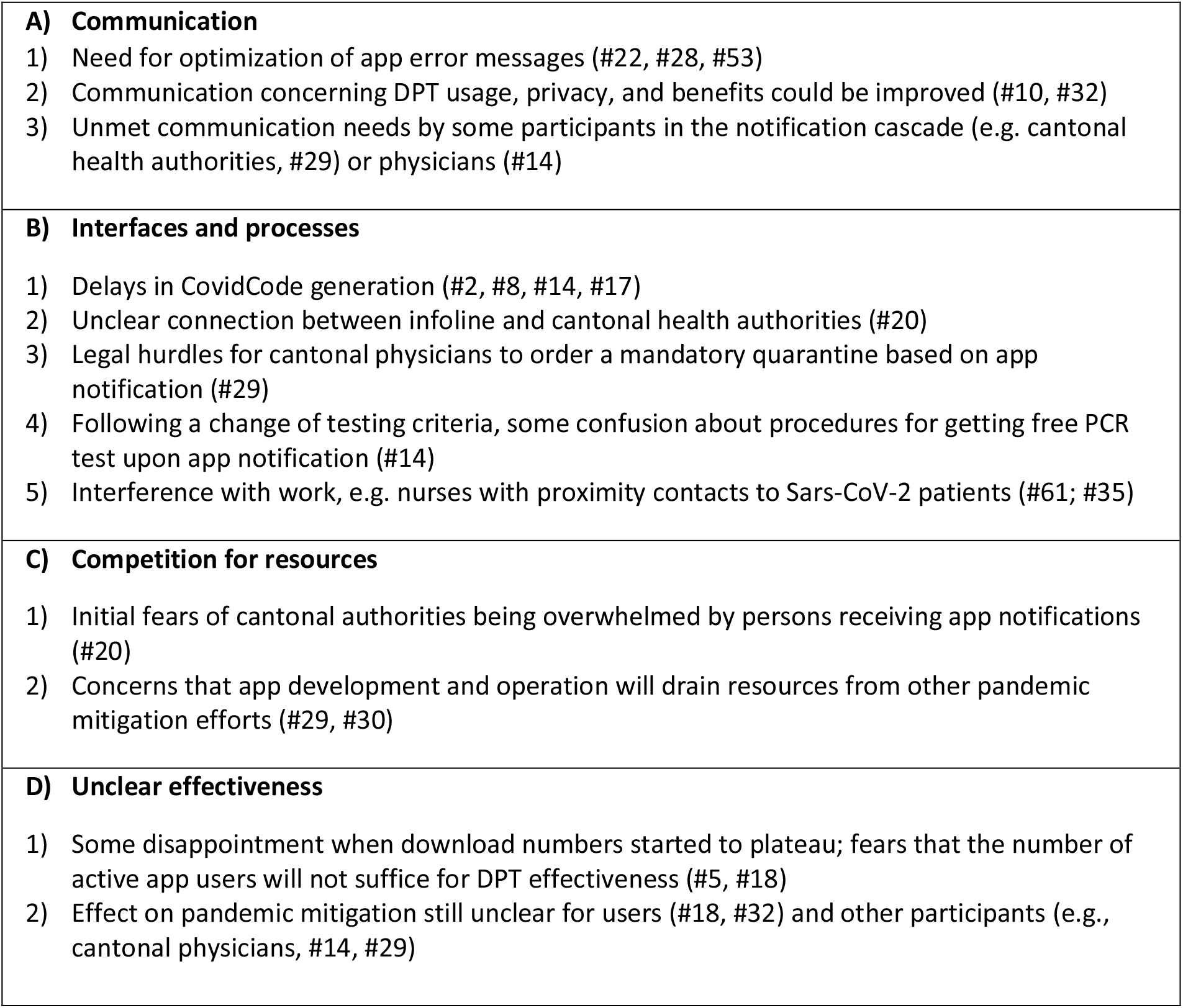
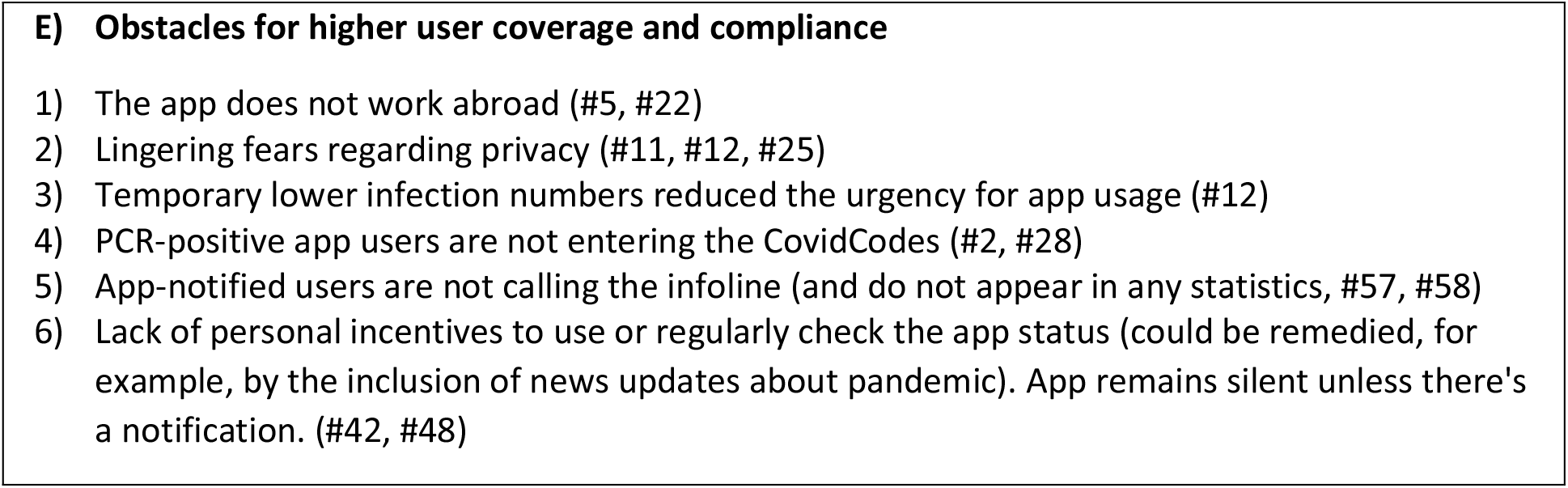
Major topics identified (#ID in parenthesis refer to individual media reports listed in supplementary table 1)

**Figure 2:**
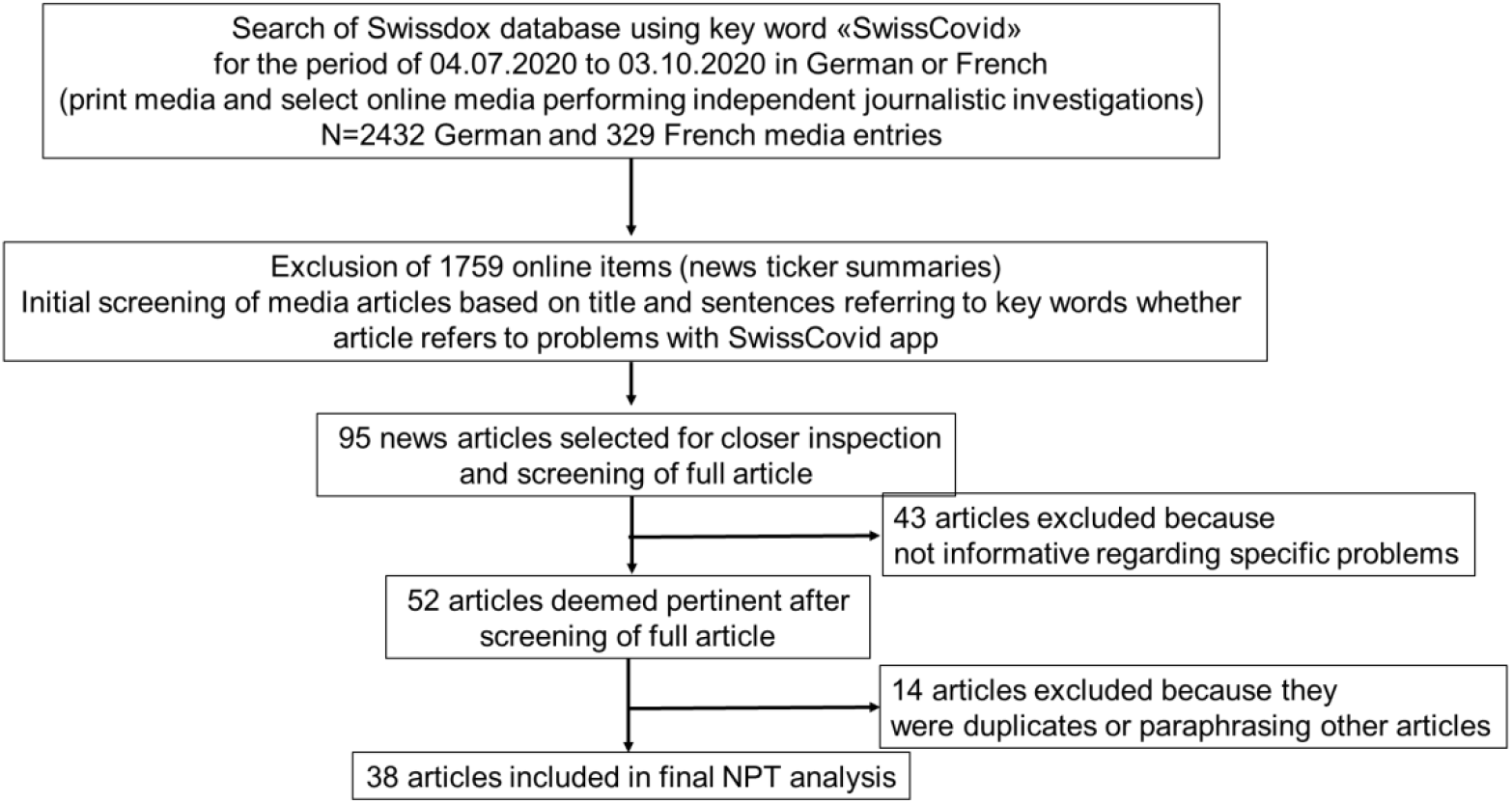
Flowchart of article selection from the Swissdox media database. Abbreviations: NPT, normalization process theory.

### Topic extraction

A more detailed analysis of the 38 pertinent articles revealed several challenges that largely fell into five topics: A) communication challenges, B) challenges to optimal DPT interfacing with other processes, C) fear of competition for limited resources with established pandemic mitigation measures, D) unclear effectiveness of DPT, and E) obstacles for higher user coverage and compliance (Table 1).

Several articles were citing communication challenges (group A, Table 1). Some reports referred to confusing app error messages, the need for intensified or improved communication to app users about the benefits and processes involved in SwissCovid, and a need for improved exchanges with other participants in the intervention, particularly the 26 cantonal health authorities. Most articles saw a solution for overcoming these challenges through intensified communication by the FOPH (being the SwissCovid product owner) with the public and other participants.

Regarding operational challenges (group B, Table 1), the most frequently echoed concern pertained to delays in sending CovidCodes to PCR-positive SwissCovid users. First media reports appeared in August 2020 after testimonies of persons who only received the codes after significant delays. Later articles also reported on procedural adjustments by cantonal health authorities to speed-up CovidCode generation and delivery.

In the context delays of CovidCode generation, other processes such as reporting of positive PCR-test results by laboratories or access to testing also came under scrutiny. Thereby, further potential problems surfaced that could affect the speed of the notification cascade. First, laboratories or physicians ordering PCR-tests may sometimes be unable to adhere to the 2-hour timeline for communication of positive test results to cantonal authorities (e.g. due to high testing volumes). Second, one physician stated that changing testing criteria and guidelines may have created some temporary confusion regarding procedures for accessing free testing by app-notified persons, which was resolved shortly after. Third, during the initial weeks after the app launch, the infoline was unable to refer app-notified callers directly to the respective cantonal health services for further evaluation. One article quoted a cantonal physician who stated data protection reasons for this referral gap. Data protection was also stated as a reason why some health authorities find it difficult to integrate DPT into manual tracing procedures. According to one cantonal physician, the (intended) inability of DPT to provide additional data on timing and place of potential exposure was diminishing its value for manual contact tracers.

Furthermore, some articles reported on the challenges for health care workers to use the app, especially when they are engaged in the care of Sars-CoV-2-infected patients. Hospitals are afraid of frequent notifications (despite personnel wearing protective gear) and ensuing confusion. Some hospitals asked their employees to switch off SwissCovid while at work.

The third group of topics (group C, Table 1) concerned the resource situation on the part of the FOPH and the cantons. A retired FOPH official and at least two cantonal physicians were cited to have some doubts regarding DPT effectiveness and were therefore concerned that DPT would compete for scarce human resources at the FOPH and cantonal health authorities. The initial referral gap between the infoline and cantonal health authorities (cf. group B) was, according to one source, driven by concerns of cantons becoming overwhelmed by app-notified contacts.

The fourth cluster of challenges (group D, Table 1) pertained to a perceived unproven DPT effectiveness. Because DPT was developed and released under immense time pressure and with limited real-world testing, doubts about the usefulness and contribution of DPT to pandemic mitigation persist. This uncertainty could potentially create a vicious cycle: the target population may not be inclined to use the app because of unproven effectiveness, but without widespread use, effectiveness cannot be demonstrated. Such concerns were echoed shortly after the public release of SwissCovid when the number of active users seemed to plateau around 1 million (on October 14, the SwissCovid app 1.67 million active users and 2.5 million downloads^12^). This perceived lack of benefit was not only confined to the public but also appeared to exist among some health authorities. Statements by two cantonal physicians are alluding to views that DPT was considered an additional burden with unclear benefits by some health authority members.

The fifth cluster of topics (group E, Table 1) was related to user coverage and compliance. In July 2020, several media reported on the plateauing (or even decreasing) user numbers, as well as on discrepancies in the numbers of generated and uploaded CovidCodes (indicating that not all PCR-positive app users chose to trigger notifications). Several explanations were explored, such as still lingering concerns about privacy (with a need for better communication), low overall case numbers of Sars-CoV-2 infections in July, or the inability to use the SwissCovid app abroad (e.g. during vacation). A frequent conclusion by the media was a need for more communication by the FOPH to address these privacy concerns and to emphasize the potential benefits of the SwissCovid app. With increasing active SwissCovid use and first manifestations of positive effects, these concerns moved somewhat to the background, but never disappeared entirely.

### Mapping of topics to NPT constructs

Table 2 illustrates the mapping of identified topics to different NPT constructs. Overall, the media analysis provided information for most of the NPT questions. Except for topic E1 (“app not working abroad”), all topics mapped well to specific NPT constructs and individual sub-questions. Ten individual topics fell into the *Coherence* construct, five into the *Cognitive Participation* construct, ten into the *Collective Action* construct, and three into the *Reflexive Monitoring* construct.

**Table 2:** Mapping of topics to Normalization Process Theory Constructs. Expressions in [] reflect adaptations to the original questions wordings [Murray et al. BMC Medicine, 2010] to better fit the current analysis.

| Normalization Process Theory Construct | Topic domains (c.f. table 1) | Assessment |
| --- | --- | --- |
| **Coherence («Making sense of the intervention»)**<br>(i.e. meaning and sense making by participants) |  |  |
| Is the intervention easy to describe | D1, D2, E3, E6 | DPT is difficult to explain, some misconceptions of what DPT should achieve (e.g. generating helpful data) or requirements for success (e.g. need of 60% participation rate in population to be successful) |
| Is it clearly distinct from other interventions | C2 | Because DPT is an adjunct to manual contact tracing, the distinction is not always obvious to all participants; DPT may even be seen as competition to manual tracing |
| Does it have a clear purpose for all relevant participants | C2, D2 | There were doubts about the purpose of DPT or even the right to co-exist with manual contact tracing |
| Do participants have a shared sense of its purpose | C2, D2 | Not all participants are convinced, including parts of the population and cantonal health authorities. |
| What benefits will the intervention bring and to whom | C1, D1, D2, E6 | Benefits are abstract, not immediately visible, and in part context dependent (e.g. role of second line of defense) |
| Are these benefits likely to be valued by potential participants | D1, D2, E2, E4, E6 | The overall potential benefits (slowing transmission) are valued by most, but doubts persist whether DPT can contribute towards that goal. |
| Will it fit with the overall goals and activity of [pandemic mitigation goals] | N/A | DPT was designed to complement manual contact tracing; In principle, DPT is well aligned with other pandemic mitigation goals. |
| **Cognitive Participation («Working out participation in the intervention»)**<br>(i.e. commitment and engagement of participants) |  |  |
| Are target user groups likely to think it is a good idea | C1, C2, D1, D2 | Some doubts seem to persist among all participants. Not all actors seem convinced of the benefits. |
| Will they see the point of the intervention easily | E3 | DPT was released during a time when case numbers were low. Benefits remained abstract and undemonstrated, in part also because of low infection numbers. Initially, this may have affected the willingness to engage in DPT work processes. |
| Will they be prepared to invest time, energy and work in it | C1, C2 | DPT was seen as competing for time and resources with other mitigation measures by some actors. Therefore, the willingness to engage in cognitive participation may have been limited. |
| **Collective Action («Executing the intervention»)**<br>(i.e. the work participants do to make the intervention function) |  |  |
| How will the intervention affect the work of [participants] | B1, B2, B3, B4, B5 | DPT introduces additional steps and processes for MCT. There were also some unclarities and frictions between different processes and interfaces (e.g. between testing labs and cantonal physicians or between PCR-positive users and cantonal physicians) |
| Will it promote or impede their work | B2, B5, C1, C2 | DPT potentially adds to the workload of MCT; app use can be problematic for health care workers |
| What effect will it have on [interactions] | B2, B3, B4, C2 | DCT notifications are an additional dimension to be covered in MCT interviews; interface between infoline and cantonal health authorities needed optimization. |
| Will staff require extensive training before they can use it | A3, B4 | In principle yes, some reports indicate an additional need for instructions or communication for some (health system) actors. |
| How compatible is it with existing work practices | C2, D2, E5, E6 | DPT is seen as something separate that adds to the workload. Data protection apparently inhibits complete DPT |
|  |  | integration into MCT. Notified users may take actions, but not always as recommended (e.g., directly seeking tests). |
| What impact will it have on division of labour, resources, power and responsibility between different professional groups? | B2, D2 | Reports indicate several «interfacing» challenges, e.g. between infoline and cantonal health authorities. Reports of confusion regarding eligibility of free PCR testing for notified users. |
| **Reflexive Monitoring («Reflecting on the intervention»)**<br>(i.e. participants reflect on or appraise the intervention) |  |  |
| How are users likely to perceive the intervention once it has been in use for a while | D1, D2 | Effectiveness still seems unclear or undemonstrated for some actors. New case numbers of Sars-CoV-2 remained relatively low for most of the observation period, thus affecting perceived effectiveness. |
| Is it likely to be perceived as advantageous for [users and other participants] | C2, D1, D2 | Seems not to be the case for all actors. |
| Will it be clear what effects the intervention has had | D1, D2 | The uncertainty regarding DPT effectiveness hampers usage and implementation. |
| Can users/participants contribute feedback about the intervention once it is in use | N/A, but indicated by some reports. | App users gave indirect feedback, e.g. via social media. Other actors had direct interactions with the FOPH and the app developers. |
| Can the intervention be adapted or improved on the basis of experience | N/A, but indicated by some reports. | App development is continuous; there is a regular exchange about possible improvements. |

The biggest concerns in the *coherence* construct pertained to unclear benefits and distinctions from other processes (especially manual contact tracing). These concerns were voiced by different participants, including app users and cantonal authorities. Besides, fears of resource competition with other mitigation measures were cited. In part, these concerns reflected the complexity of the SwissCovid app notification cascade, and possibly also an incomplete understanding of the role requirements by some participants.

Multiple reports also indicated challenges in the *cognitive participation* domain, which overlap with other NPT constructs. Specifically, the interplay between different actors may require optimization, as well as clarifications regarding resource situations (e.g., on the part of cantonal authorities).

Furthermore, many of the reported problems in the *collective action* domain indicated a need for optimized integration of SwissCovid into existing work processes. From the viewpoint of some cantonal health authorities, SwissCovid was perceived to add to the existing workload, but with unclear benefits. Additionally, some statements reflected a need for additional communication and knowledge transfers to different participants, for which the FOPH was seen in the lead.

By contrast, there were fewer media reports regarding the *Reflexive Monitoring* construct. But some reports indicate that communication between different participants (FOPH, cantonal physicians, infoline operators) is ongoing, and optimizations have been planned or even put in place (e.g., better coordination between infoline and cantonal health authorities; integration of news into SwissCovid app to provide additional user incentives).

## Discussion

DPT is a complex public health intervention to mitigate the Sars-CoV-2 pandemic. Its effectiveness depends on appropriate embedding into a country’s overall TTIQ strategy and requires multiple, timely actions by different actors. This article describes early experiences with DPT implementation in Switzerland, which has one of the longest track records of decentralized DPT operation.

Based on Swiss media articles published during the first three months after DPT release, this study presents different challenges in non-technical DPT implementation. These challenges included unclear DPT benefits, which affected commitment and raised fears among different health system actors for resource competition with established pandemic mitigation measures. Moreover, media reports indicated process interface challenges in the notification cascade (e.g., in the hand-over of app notified users from the infoline to responsible cantonal authorities), as well as misunderstandings and unmet communication needs on the side of some health system actors. Finally, some reports suggested misaligned incentives, both for app usage by the public as well as for process engagement by other actors in the app notification cascade.^13^

In the Sars-CoV-2 pandemic, timely diagnosis,isolation and quarantine are essential processes, which DPT apps intend to support. However, procedural frictions can lead to delays, which also affect DPT effectiveness. Examples are late deliveries of CovidCodes (codes to trigger notifications) or uncertainties regarding access to PCR-tests upon app notification. Furthermore, most challenges identified in the media search bear close relations to constructs of NPT. Specifically, many identified challenges mapped well to the *coherence* (“making sense of the intervention”) and *Collective Action* (“executing the intervention”) domains. For example, unmet communication and training needs regarding DPT usefulness and integration into existing workflows seemed to exist, which hindered stakeholder onboarding and optimal process flows in the notification cascade. Perceived lack of usefulness is also affecting the uptake of the DPT app in the population. However, “sense-making” by different participants may also be context-and time-dependent. The SwissCovid app was released at a time of relatively low Sars-CoV-2 incidence, which meant that DPT effects could not become apparent immediately.^14^ But in late August 2020, the FOPH presented data indicating rising numbers of app users who sought PCR-testing after a SwissCovid notification and then tested positive, which may have alleviated some of the concerns regarding effectiveness.

The present analysis may contribute to the international debate on DPT on two levels. First, it provides insights into challenges of DPT implementation from a country with one of the longest track records of DPT implementation and a complex, federalistic health system. Second, by applying the NPT framework to classify different reports, the analysis also contributes to a methodological level by illustrating the usefulness of the NPT approach. Ideally, NPT or similar implementation frameworks should already be considered during the development and release of novel technologies, which was hindered by immense time pressures by the pandemic situation.

Second, many of the challenges highlighted by the media were well reflected by the NPT constructs. NPT postulates that individual and collective actions need to be in synchronization for a complex intervention to be successful. To quote one of the foundational papers of NPT: “The starting point of the theory is that to understand the embedding of a practice we must look at what people actually do and how they work.”^15^ The present analysis indicates that NPT can indeed provide a useful framework to classify DPT challenges and may help to identify suitable optimization. For example, as suggested by the *reflexive monitoring* construct, some media reports indicate continuous DPT process adaptations, as well as constant communication with actors and assessments of potential improvements.

Some limitations of the study and its interpretation are worth noting. The reliance on published media reports (and not, for example, on stakeholder interviews) may have limited the diversity and level of detail of the current debates about DPT. It may also have missed aspects that were never raised by the media but discussed bilaterally between different actors.The reliance on published media report was an advantage as it enhanced the reproducibility of our study. Furthermore, it should be noted that the challenges outlined in this report do not reflect the status quo situation, and it is not intended to pass judgment on the success of DPT or the roles of the different participants in Switzerland.

To summarize, the analysis of media reports on implementation challenges for DPT in Switzerland demonstrates that the non-technical implementation of DPT must not be forgotten. The experiences in Switzerland indicate that the technical aspects work well, but in some instances, the non-technical processes led to bottlenecks in the notification chain. This is understandable given the multiple interactions required between different participants. The lessons from Switzerland, one of the earliest adopters of DPT, and the demonstration of the usefulness of NPT for planning and analyzing NPT implementation may hopefully serve as an inspiration for other countries developing their own DPT implementation.

## Data Availability

Data are available from the corresponding author upon request.

---

**Supplementary table 1.**

| File # | Title | Date | Medium |
| --- | --- | --- | --- |
| 1 | «Wir haben viel Grund zur Hoffnung» | 27.09.2020 | SonntagsZeitung |
| 2 | Corona-App erhält Daten oft zu spät | 14.08.2020 | NZZ |
| 3 | So ist die SwissCovid-App komplett nutzlos | 14.08.2020 | NZZ |
| 4 | Digitale Verwaltung braucht Transparenz | 23.07.2020 | NZZ |
| 5 | Geht der SwissCovid App die Luft aus? | 16.07.2020 | NZZ |
| 6 | «Es muss jemand auf den Tisch hauen» | 28.08.2020 | TagesAnzeiger |
| 7 | SP fordert dringend Verbesserungen beim Contact-Tracing | 28.08.2020 | Aargauer Zeitung |
| 8 | Kanton verschickt am Sonntag keine Covid-Codes | 27.08.2020 | Aargauer Zeitung |
| 10 | Epidemiologe kritisiert Behörden | 23.07.2020 | NZZ |
| 11 | Noch ein Rückschlag für die Corona-App | 21.07.2020 | Aargauer Zeitung |
| 12 | Siebzig Covid-Meldungen in sieben Tagen | 14.07.2020 | Aargauer Zeitung |
| 14 | Ärgerliche Corona-Test-Odyssee | 06.08.2020 | regio.ch |
| 15 | «Generelles Unbehagen» gegen Covid-App | 29.08.2020 | Bieler Tagblatt |
| 17 | Die Corona-App bleibt stumm | 20.08.2020 | St. Galler zeitung |
| 18 | Zu wenig Anrufer: Bund weiss noch nicht, ob Corona-App hilft | 16.07.2020 | blick.ch |
| 19 | Warum versagen die Kantone bei der SwissCovid-App? Es gibt einen bösen Verdacht | 28.08.2020 | watson.ch |
| 20 | Covid-App-Frust und ein wenig Hoffnung: So lief die Corona-Konferenz in Bern | 19.08.2020 | watson.ch |
| 22 | Das sind die grössten «Baustellen» bei der SwissCovid App | 07.07.2020 | watson.ch |
| 25 | «La Suisse n'était pas préparée à la bonne crise» | 28.07.2020 | le temps |
| 27 | Une application qui sauve des vies | 29.08.2020 | le temps |
| 28 | L'utilité de l'app SwissCovid se dessine | 29.08.2020 | le temps |
| 29 | Alarm ins Nirgendwo | 27.08.2020 | WOZ |
| 30 | Daniel Koch bot mehrmals seinen Rücktritt aus der Taskforce an | 12.09.2020 | TagesAnzeiger |
| 32 | Wie das BAG die SwissCovid-Nutzung ankurbeln will | 28.08.2020 | netzwoche.ch |
| 33 | SwissCovid: App-Entwickler Mathias Wellig über Tücken des Contact-Tracings | 27.08.2020 | netzwoche.ch |
| 35 | Corona-Fehlalarme bei Pflegepersonal | 05.09.2020 | Basler Zeitung |
| 36 | Schwarzpeterspiel um Corona Tracing | 29.08.2020 | Blick |
| 37 | Erstmals unerwartete Coronafälle entdeckt: "Wir haben den Sinn der App bewiesen" | 28.08.2020 | Aargauer Zeitung |
| 39 | Von wegen «ein Witz»: So reagieren die Kantone auf die harte Corona App-Kritik | 20.08.2020 | watson.ch |
| 42 | Schweizer vertrauen der Swiss-Covid App mehr als den Onlineshops | 21.08.2020 | 20 minuten online |
| 48 | Die Corona-App bringt mich um den Schlaf | 17.07.2020 | Basler Zeitung |
| 50 | Die Swiss-Covid-App alarmiert nicht immer | 26.07.2020 | Sonntagszeitung |
| 52 | Der Funke will nicht springen | 23.07.2020 | Weltwoche |
| 53 | Sandro Brotz motzt gegen Swiss-Covid-App | 22.07.2020 | blick.ch |
| 57 | Gewarnte gehen nicht über Infoline | 17.07.2020 | TagesAnzeiger |
| 58 | Wirkung der Swiss-Covid-App kann noch nicht genau gemessen werden | 16.07.2020 | Basler Zeitung |
| 60 | Ringen um Akzeptanz und Wirksamkeit | 11.07.2020 | Schaffhauser<br>Nachrichten |
| 61 | Angst vor zu vielen Fehlalarmen | 09.07.2020 | Linth-Zeitung |

